# Increasing influenza vaccination rates among care home staff: Economic evaluation of the FluCare intervention within a cluster-RCT

**DOI:** 10.64898/2026.06.06.26355050

**Authors:** Adam P. Wagner, Helen Risebro, Allan Clark, Susan Stirling, Erika J. Sims, Wasim Baqir, Veronica Bion, Jeanette Blacklock, Linda Birt, Robert Bryant, Luke Cook, Tony Dean, Alys Wyn Griffiths, Cecile Guillard, Richard Holland, Andy Peter Jones, Liz Jones, Thando Katangwe-Chigamba, Jennifer Pitcher, Sion Scott, David J. Wright, Amrish Patel

**Affiliations:** Health Economics Group, Norwich Medical School, University of East Anglia, Norwich, UK; National Institute of Health and Care Research (NIHR) Applied Research Collaboration (ARC) East of England, Cambridge, UK; Norwich Medical School, University of East Anglia, Norwich, UK; Norwich Clinical Trials Unit, University of East Anglia, Norwich, UK; Pharmacy Integration Fund, NHS England, London, UK; School of Healthcare, University of Leicester, Leicester, UK; Public contributor and Lay Advisory Group (LAG) member, Liverpool, UK; Askham Village Community, Doddington, UK; Norfolk Local Pharmaceutical Committee (LPC), Great Bircham, UK; School of Medicine and Population Health, University of Sheffield, Sheffield, UK; University of Exeter Medical School, University of Exeter, Exeter, UK; Centre for Health Services Studies, University of Kent, Canterbury, UK; National Care Forum, Coventry, UK; School of Economics, University of East Anglia, Norwich, UK

## Abstract

**Introduction:** Care home (CH) influenza vaccination of staff improves resident health, yet uptake remains low at just over 11% (England, 2025/2026). We report an economic evaluation (EE) of ‘FluCare’, an intervention to increase staff influenza vaccination through: vaccination clinics at CHs; promotional materials; and CH financial incentives.

**Method:** Seventy-five CHs were randomised to FluCare or control. A cost-consequence analysis took the influenza vaccination programme funder perspective, but also extended to the National Health Service (NHS) and CH perspective. Costs included: influenza vaccination; administration fee; FluCare components; CH resident NHS utilisation. Outcomes were: staff influenza vaccination rates; staff sickness; and resident mortality. Sensitivity analyses excluded intervention CHs that did not host vaccination clinics.

**Results:** Compared to control CHs, adjusted analysis found intervention homes with a mean absolute increase in vaccination rates of 1.8% (95% CI: -6.0%, 10.8%; p=0.572) at an increased cost of £451 (95% CI: £239, £675; p<0.001) to the vaccination programme funders: £249 per additional percentage point (PAPP) per CH. Vaccination clinics were delivered late in the influenza season, with 80% taking place from February 2023.

Including only intervention CHs that hosted staff flu vaccination clinics (23/35), increases the mean difference to 10.1% (95% CI: 0.9%, 21.9%; p=0.018) and costs to £805 (95% CI: £603, £1,079; p<0.001): £79 PAPP per CH. Differences between trial arms in other costs and outcomes were marginal and generally non-significant.

**Conclusions:** FluCare delivered little improvement when staff flu vaccination clinics did not occur and had little impact on other costs/outcomes. Cost-effectiveness depends on willingness-to-pay for increased staff vaccination, but cost PAPP per CH improved from £249 to £79 when only CHs hosting clinics were considered. Late implementation, likely reduced impact by limiting clinic delivery, as reflected in sensitivity analysis. Future evaluations should implement FluCare earlier in the season.

**Trial registration:** ISRCTN22729870. Registered on 24 August 22. Secondary identifiers: R209939, IRAS 316820, CPMS 53812.

## Introduction

During the 2022/2023 influenza (flu) season, UK flu-related mortality was estimated at almost 15,000, with 86% of deaths occurring among those aged ≥65 [1]. Older adults are at increased risk of flu-related morbidity and mortality [2], with underlying chronic conditions exacerbating disease among care home (CH) residents [3]. The CH environment itself exacerbates risk of flu outbreaks, given prolonged close contact between residents and staff contributing to rapid disease spread [4].

Vaccine response among older adults is generally lower [5] impacting negatively on economic benefits of directly vaccinating over-65s [6]. To better protect residents, attention has shifted toward vaccinating CH staff, which has been shown to improve resident outcomes [7][8][9]. In the UK, influenza vaccination is freely available to CH staff [10], accessible off-site in their own time. However, despite recent policies promoting vaccination [11][12][13], only 11.3% of total staff of older adult CHs in England received a flu vaccination in the 2025/26 season [14].

A multi-component intervention called ‘FluCare’ has been developed using behavioural science to underpin its design [15][16]. Its components tackle access, cost, and cognitive beliefs, to support behaviour change. A key component is on-site staff flu vaccination clinics in CHs delivered by vaccine providers (VPs) to staff free of charge, supported by financial incentives for both VPs and CHs to improve vaccine accessibility and uptake.

While there are general economic explorations of the impact of flu vaccinations (e.g. [6]), fewer focus on workplace interventions targeting UK health and social care workers [17][18]. Among the relatively few interventions focused on CH staff [8][19][20][21][22], we are unaware of economic evaluations of these interventions – our study presents unique data. Hayward et al. [8] considers the impact on health care resource use (hospital admission and GP consultations) by residents, but they do not cost this resource use, nor are intervention delivery costs reported. In addition, previous studies have given limited attention to outcomes relevant to CH providers, such as staff sickness absence and agency staff use, despite evidence that staff vaccination may reduce absenteeism and associated costs [23, 24].

We report a trial-based economic evaluation (EE) of FluCare, using data collected within a two-arm, cluster randomised controlled trial (cRCT) evaluating its effectiveness at improving uptake of flu vaccination by CH staff in England, compared to usual care. We undertake a cost-consequence analysis, focusing on the perspective of the vaccination funder. This is extended to consider impacts on the NHS and CHs, taking note of NHS use by residents and costs of staff sick days.

## Materials and Methods

### Trial design

FluCare was evaluated within a two-arm cRCT. A feasibility study [15] refined study design informing data collection methods and selection of outcome measures, including the primary outcome: percentage of staff vaccinated. The project’s lay advisory group (LAG) consisted of public members with CH experience. LAG members were part of project management group meetings and actively involved throughout the project, contributing to reviewing materials including co-authorship of this manuscript, and day-to-day project delivery. Intervention development, cRCT protocol, cRCT clinical effectiveness results and cRCT process evaluation are reported elsewhere [15][25][16][26]. A signed Health Economic Analysis Plan (HEAP), finalised prior to analysis, is available from APW (HEAP deviations noted in S1 Text).

Randomisation was performed by data management within the Norwich Clinical Trials Unit. CHs were allocated electronically using block randomisation, stratified by percentage of staff identifying as white. Recruitment and CH facing teams had no access to the allocation sequence [25], but it was otherwise unfeasible to further blind the research team.

Given FluCare may impact multiple domains and perspectives (e.g. staff, CHs, and NHS), we use a cost-consequences analysis (CCA) for a more holistic evaluation than, for example, a cost-effectiveness analysis focusing on one particular outcome [27].

### Ethics approval and consent to participate

Ethical approval was received from the University of East Anglia’s (UEA’s) Faculty of Medicine and Health Sciences Research Ethics Subcommittee on 01/08/2022. Study approval number: ETH2122-2419. An ethics amendment to extend recruitment to 30/11/22 was approved by the same committee on 07/10/22. Study approval number: ETH2223-0272 (amendments). Governance approval was received from the UK Health Research Authority on 15/08/2022 (study approval number IRAS 316820).

Written informed consent to participate in the study was obtained from care home managers and vaccination providers. No identifying images or other personal or clinical details of participants are presented here or will be presented in trial reports.

### Population

The cRCT took place within CHs across England during the 2022/2023 flu season (CH recruitment opened 1^st^ September 2022, with resident health log data collection spanning 31^st^ October 2022 to 12^th^ April 2023). CHs were community-based, and either local authority, charity, private or corporate managed. They were required to be registered with the Care Quality Commission (CQC) to provide care for older adults (65 or older; but could offer residential, nursing, or dementia care), employed ten or more staff, and self-reported staff flu vaccination rates below 40% during flu season 2021/2022. Within CHs, the primary focus was staff (including agency staff and volunteers), but aggregate (anonymised) information was also collected about residents. CHs were ineligible if they participated in: other behaviour change interventions; or the feasibility study [15].

### Sample size

A total of 75 CHs were randomised (37 to intervention; 38 to control). CHs missing either or both ‘logs’ (data collection tools – see: Outcomes) were excluded: 5 CHs had both logs missing (2 intervention; 3 control); a further 3 control CHs had no resident health logs. Consequently, in the base-case EE analysis, having applied steps to address missing data (see: Missing data), we included 67 CHs (35 intervention; 32 control) with complete data on: i) influenza vaccine programme funder resources/costs; and ii) resident NHS use. For the primary outcome analysis, a sample size calculation estimated a requirement of 31 CHs per trial arm [25]. See the clinical effectiveness paper’s [16] CONSORT diagram for further detail.

### Comparator and intervention

#### Comparator

The control arm consisted of usual care, augmented with monthly data collection focusing on staff vaccination and resident NHS use (see: Outcomes). CH staff were free to access usual sources of flu vaccination and providers of these vaccinations were able to reclaim the vaccine cost plus an administration fee from NHS England. CH managers/owners were aware of their CH’s participation, but no additional information or materials were provided. Monthly outcome data was requested by the research team to improve data quality, with follow-up and feedback to CHs on identified issues in data logs. The prior feasibility study evidenced no reactivity bias from such monitoring [15].

#### Flu-Care intervention

Beyond usual care noted above, the multi-component ‘FluCare’ intervention comprised: promotional materials for each CH; CH incentive scheme; and provision of vaccination clinics hosted by CHs.

Promotional materials were co-produced and comprised: a short video of stakeholders (GP, pharmacist, CH manager, residents, and CH staff) promoting vaccination benefits; and supporting materials (including posters and leaflets).

FluCare offered CHs an £850 incentive payment of £850 where ≥70% of staff were reported vaccinated. Financial incentives have previously been used to promote staff flu vaccination uptake in NHS hospitals [28] and in long term care facilities in the US [29]; furthermore, many local authorities pay premia to CHs to incentivise care quality in general [30]. Use of incentive payments was viewed as particularly powerful by sector leaders as it signalled equity between health (NHS) and social care.

FluCare was designed to offer all CH staff, including administrative, kitchen, agency staff and volunteers, a free vaccine at FluCare vaccination clinics hosted by their CH. Providing vaccination on-site reduces known barriers such as travel, time and loss of non-work hours [31].

Vaccination clinics were delivered by community pharmacies (often local high street or retail pharmacies) or GP surgeries, although clinic delivery personnel extended beyond GPs or pharmacists (see S16 Table for full range). Clinics were delivered at date/times agreed between VP and CH. In the UK, operational and financial pressures can limit both groups’ ability to provide vaccination clinics in care homes. Community pharmacists must maintain a pharmacist on site to keep pharmacies open [32], while GPs are typically only reimbursed for vaccinating staff registered with their practice [33]. FluCare reduces these barriers by guaranteeing a minimum income for VPs delivering clinics in CHs. VPs received payment for delivering up to four clinics, with a minimum fee of £300 per clinic (detail about clinics, including duration, are provided in S5 Text). For pharmacy VPs, the £300 fee was reduced by income accrued from the NHS Business Services Authority (NHSBSA) administration payment. In principle, GP practices could also claim this administration fee; however, in practice did not given the significant burden in registering each CH staff member (if not already registered) with the practice (see Table 1).

**Table 1:**
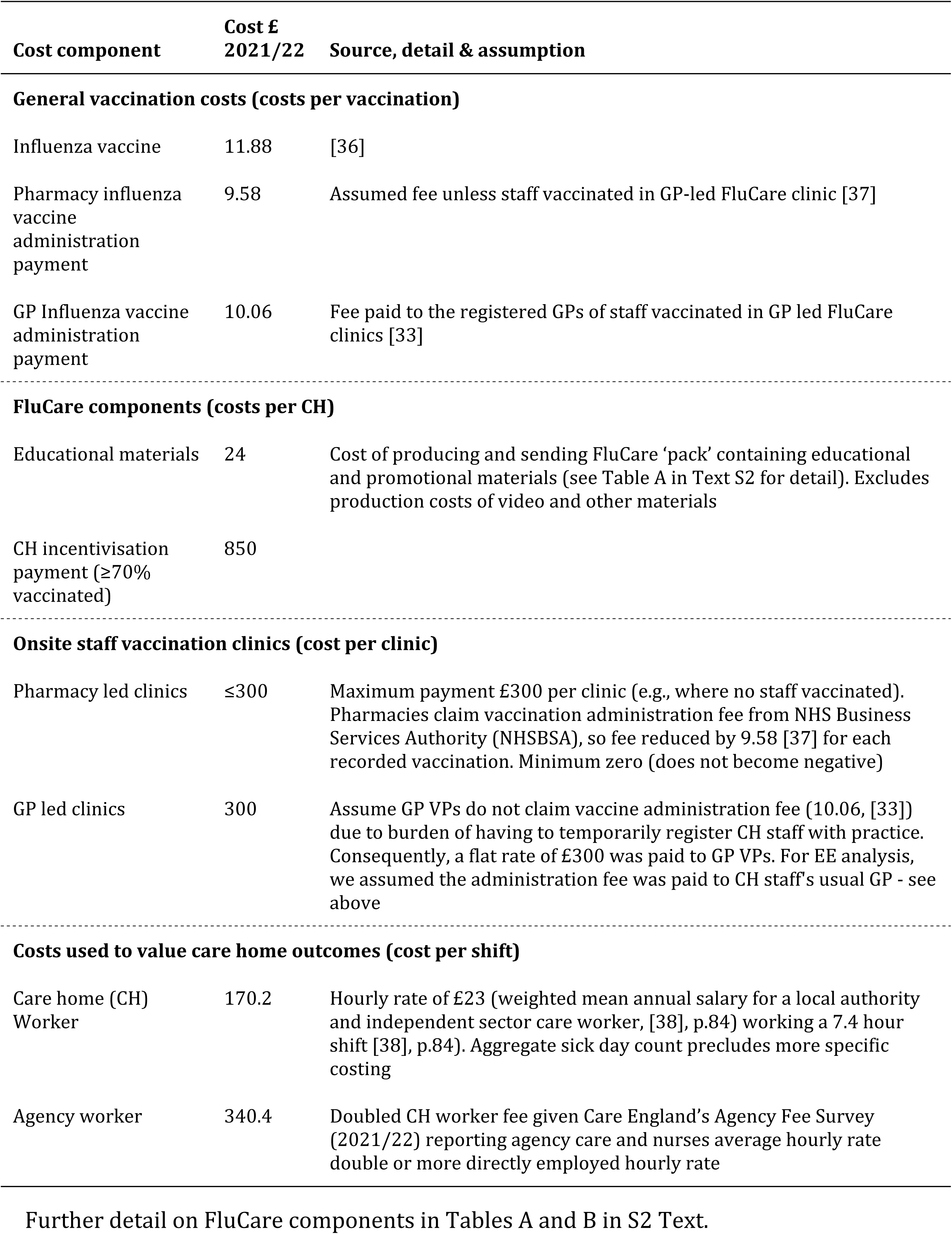
Unit costs (£, 2021/2022) for vaccination and FluCare components, with detail of sources and assumptions.

While performance monitoring with feedback was intended to be conducted, this was not feasible within the trial’s available resources; thus, was undelivered, and so is excluded from costings. Further information on intervention development can be found in [34], and a Template for Intervention Description and Replication (TIDieR; [35]) was published with the clinical effectiveness paper [16].

### Time horizon and perspective

A time horizon of approximately five months was used, corresponding to trial length. The primary perspective was that of the flu vaccination programme funder. We also adopt a wider perspective to include impacts to the UK health service (labelled as ‘NHS perspective’), incorporating both the i) National Health Service (NHS) funding the vaccination programme *and* ii) use of NHS by CH residents; associated results are reported in supplementary materials (table B in S2 Text, and S3-S16 Tables).

### Outcomes

Consistent with a CCA, we consider multiple outcomes [39][27]. Unless otherwise noted, the unit of observation is CH.

The key outcome was percentage of staff vaccinated, where for a given CH this is the total number of staff vaccinated in the flu season (September 2022 to end April 2023) over total number of staff employed at any point during the flu season. We define staff to include: directly employed; bank; agency; and volunteers. Other staff-related outcomes include totals of: sick days; shifts worked by agency staff; and number of agency staff utilised. Source data for these outcomes were collected from a ‘staff log’, a spreadsheet completed (generally) monthly by each CH, recording demographics, job details and vaccination status for each staff member. Within these logs, CHs also provided an aggregate total count of staff sick days.

A ‘resident health log’ – again, an Excel spreadsheet completed (generally) monthly by each CH – provided aggregate information on use of NHS services and resident deaths. These included hospital use (planned and emergency attendance and admissions), GP and nurse consultations, as well as paramedic attendances.

‘Clinic logs’ completed by vaccination providers recorded the: job role of the person delivering the clinic; number of vaccinations delivered; time to travel, organise and deliver clinics.

Further data collection detail is in the HEAP and clinical effectiveness paper [16]. Care home characteristics [including: resident numbers; home type (residential/nursing/both)] were collected at baseline using a ‘Site Profile Questionnaire’, completed by CH managers or designated staff.

### Measuring resource use and costs

Uptake of influenza vaccination was based on the vaccination statuses recorded in staff logs and costed using unit costs in Table 1. We assumed CH staff were vaccinated at a pharmacy, unless indicated as vaccinated at a FluCare clinic; if the FluCare clinic was GP-led, we assumed the staff member’s registered GP practice claimed their administration fee.

Resources and associated costs of the FluCare components are summarised in Table 1, with further detail given in S2 Text. Costs of vaccination clinics hosted by CHs varied with the type of VP managing them (GP or pharmacy-led), number of clinics delivered, and the staff vaccinated (impacting pharmacy led clinics) – source information was taken from clinic logs, with a log completed per clinic. Where onsite clinics were delivered, all clinics within a given CH were either pharmacy- or GP-led. Intervention costing focused on ongoing delivery costs (e.g., excludes online-video development costs).

For the vaccination funder perspective, total costs for each CH were calculated by summing vaccination costs for its staff, along with costs of FluCare components for CHs in the intervention arm.

The NHS perspective extended the vaccination funder perspective to include residents’ use of NHS services. For each CH, we had aggregate data about a range of NHS use by residents – associated unit costs can be found in table B in S2 Text.

Costs are reported in £ in 2021/2022 values. Given the time horizon of less than a year, no discounting is applied.

### Missing data

Missingness was low, but where it occurred, it typically related to CHs returning no logs or incomplete logs. CHs failing to return required logs (staff and resident health) were excluded – reducing the sample to 35 (95%) intervention and 32 (84%) control CHs. Among these 67 CHs, we assumed a value of zero where count-based or binary outcomes were missing data. Impact of the latter is explored in a sensitivity analysis (See Sensitivity analyses section). These exclusions and imputation approach accord with the statistical analysis plan and clinical effectiveness paper [16].

### Data analysis

The primary observational unit is CH. Data collection ran from late October 2022 until mid-April 2023, with CHs providing data spanning different durations.

One set of comparison between arms compared outcomes (sick days, agency shifts, agency staff counts, resident deaths) and NHS resource use in terms of annual rates per staff member or resident, depending on the outcome. Calculated for each CH, these were standardised for different numbers of staff/residents and periods of data collection. CH outcome rates between arms were compared using two-sample t-tests. Due to low numbers, resident deaths are presented as rates per 100 residents.

Another set of comparisons applied to both these outcomes and costs, compared CHs across the flu season (September 2022 to end April 2023). In unadjusted analysis, two-sample t-tests were used to compare arms (e.g., for staff sickness totals, each CH has a season total, and these totals were compared using two sample t-tests). In the adjusted analysis, for each variable of interest, a suitable regression model was fitted, with an arm covariate estimating differences between arms. Each regression model included covariates to adjust for stratification and confounding by the following factors: strata (above/below 23% of staff identifying as non-white); type of CH (residential/nursing/both); size of CH (either number of resident beds or staff employed, depending on outcome); and duration of data collection. Confidence intervals (CIs) and associated p-values were estimated using non-parametric bootstrapping (see Identifying sample uncertainty section). Poisson models were used for count-based variables and linear regressions otherwise. Differences in counts between arms in Poisson models were estimated using average marginal effects (AMEs; using the margins package [40]).

From the prior unadjusted/adjusted analysis, we estimated, per CH, incremental costs and incremental differences in percentages of staff vaccinated. These incremental estimates were then divided to calculate the incremental cost-effectiveness ratios (ICERs) which gave the additional cost per percentage of vaccination per CH. We also report this ICER scaled by the mean CH staff complement (mean=63) to give the cost per additional staff member vaccinated per CH.

### Identifying sample uncertainty

Sample uncertainty was estimated using stratified bootstrap resampling: each bootstrap randomly selects, within strata and arm of the trial with replacement, a subset of CHs. Within each bootstrap sample, for each variable of interest, a suitable regression model was fitted (see Data analysis section), with a covariate for arm to estimate adjusted differences between arms (in Poisson models, we used adjusted AMEs). We use 1,000 bootstrap samples, in line with suggestions from Efron & Tibshirani [41] when estimating CIs using the percentile method (e.g. using 2.5 and 97.5 percentiles to estimate 95% CIs).

We considered uncertainty in mean incremental costs (for both influenza vaccine programme funder and NHS perspectives) and mean incremental differences in proportion of staff per CH vaccinated. From the bootstrap replicates, cost-effectiveness planes were constructed for the base-case and key scenarios; each plane displays the estimated mean incremental cost and mean incremental difference in percentage of staff vaccinated for each replicate, with the location and spread of points showing uncertainty [42]. Additionally, cost-effectiveness acceptability curves (CEACs) were constructed for the base-case and key scenarios, showing the probability of FluCare being cost-effective compared with usual care at a range of willingness-to-pay (WTP) thresholds for each additional percentage point of vaccination per CH [43].

### Sensitivity analyses

Three sensitivity analyses were conducted. The first addressed a key interest about the impact of including only intervention CHs hosting a FluCare vaccination clinic (thus excluding intervention CHs that had no onsite FluCare clinics). Second, we excluded CHs with incomplete logs (see Missing data section). Third, we explored the impact on the base-case and the first sensitivity analysis of excluding: i) agency staff and volunteers; ii) : agency, volunteers and bank staff.

### Secondary analyses

These explored:

- Estimating the value to CH of impacts of influenza vaccinations on staff sick days and utilisation of agency staff. We valued these as in Table 1, in two ways: i) loss of productivity of one staff member; and ii) replacing that staff member with another.
- Valuing mortality differences between arms as crude quality adjusted life years (QALYs). We assumed the utility of residents in CHs based on a previous trial [44] – for further detail see Tables A and B in S3 Text.
- Impact of analysing cost-effectiveness using staff level effectiveness data – see Table A in S4 Text.
- Costing FluCare vaccination clinics based on timings recorded on clinic logs, using staff costs sourced from [38] – see Tables A and B in S5 Text. This contrasts with the set fee-based costing approach used in our main analysis (c.f., Table 1).

## Results

### CH characteristics

During the 22/23 flu season, 75 care homes were randomised to intervention and control, two or more months into the season. A Consort diagram is provided in Figure 1. After excluding CHs that did not return logs, and imputing missing data in logs of the remaining CHs (see Missing data section), 35 and 32 were included in intervention and control arms respectively in the base-case (89%, 67/75). Characteristics of these homes are summarised in Table 2: proportionately fewer CHs in the intervention arm were exclusively nursing, and most homes across both arms were privately owned. On average, intervention homes had fewer residents and staff members. Distributions of staff types were similar.

**Fig 1:**
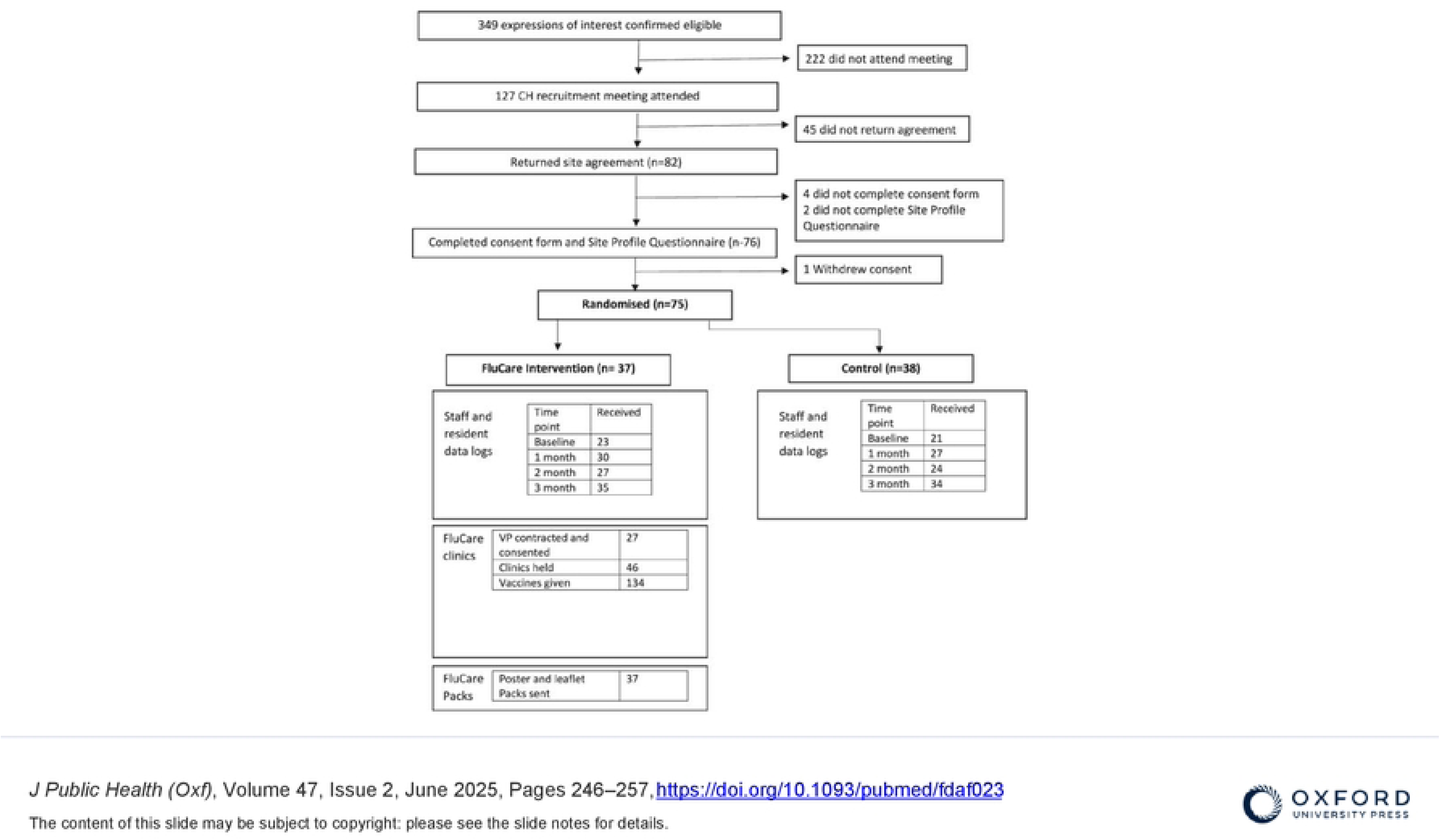
Consort diagram.

**Table 2:** Description of care homes included within base-case analysis (complete intervention costs and NHS use)

|  | Level/statistic | Int | Cont |
| --- | --- | --- | --- |
| N |  | 35 | 32 |
| Registration | % residential | 57 | 38 |
|  | % nursing | 0 | 16 |
|  | % both | 43 | 47 |
| Ownership | % local authority | 3 | 0 |
|  | % charity | 6 | 28 |
|  | % private | 91 | 72 |
| Ethnicity strata | % non-white staff<br>≥23% | 54 | 56 |
| Residents | Mean | 38 | 43 |
| Staff | Mean | 55 | 72 |
| Total staff | N | 1,929 | 2,305 |
| Directly employed | % | 88.4 | 91.0 |
| Bank | % | 10.0 | 8.1 |
| Agency | % | 1.1 | 0.7 |
| Voluntary | % | 0.5 | 0.2 |

Int=intervention. Cont=control. Diff=difference. CI=confidence interval

### Costs

Costs between arms for the base-case analysis are reported in Table 3.

**Table 3:** Vaccination and intervention costs/rates per CH per flu season (base-case with complete intervention costs and complete NHS use)

| Cost (£, 21/22)/<br>outcome | CHs (N) |  | Unadjusted per home per flu season |  |  |  |  |  | Adj. per home per flu season |  |  |  |
| --- | --- | --- | --- | --- | --- | --- | --- | --- | --- | --- | --- | --- |
|  | Int | Cont | Int | Cont | Diff. | 95% CI |  | p | Adj.<br>diff. | 95% CI<sup>†</sup> |  | p<sup>†</sup> |
| Vaccine* | 35 | 32 | 173 | 215 | -42 | -128 | 43 | 0.326 | 14 | -44 | 81 | 0.598 |
| Vacc. admin fee* | 35 | 32 | 140 | 174 | -33 | -102 | 35 | 0.335 | 12 | -35 | 66 | 0.586 |
| Promotion | 35 | 32 | 24 | 0 |  |  |  |  |  |  |  |  |
| Clinic fees | 35 | 32 | 370 | 0 |  |  |  |  |  |  |  |  |
| Incentives | 35 | 32 | 49 | 0 |  |  |  |  |  |  |  |  |
| Total* | 35 | 32 | 756 | 389 | 367 | 127 | 606 | 0.003 | 451 | 239 | 675 | <0.001 |
| % vaccinated* | 35 | 32 | 28.3 | 27.0 | 1.3 | -8.6 | 11.3 | 0.788 | 1.8 | -6.0 | 10.8 | 0.572 |
| Funder ICER: £/% |  |  |  |  | 273 |  |  |  | 249 |  |  |  |
| Funder ICER:<br>£/person |  |  |  |  | 435 |  |  |  | 397 |  |  |  |

CHs=care homes. Int=intervention. Cont=control. Diff=difference. CI=confidence interval. Adj. diff.=adjusted difference. ICER=incremental cost-effectiveness ratio
Costs/rates by: i) unadjusted rates per home per flu season; ii) adjusted rates per home per flu season [controlling: for home type; randomisation strata; duration of data collection; and staff numbers (\*)].
<sup>†</sup>Confidence intervals and p-values derived from bootstrap resampling (n=1,000) – see ‘Identifying sample uncertainty’.

In the flu vaccination programme funder perspective, compared to the control arm, adjusted total costs were significantly (p<0.001) higher by £451 (95% CI: £239, £675) for intervention homes. The most costly FluCare component was provision of vaccination clinics, held between November 2022 and April 2023 (with 80% taking place from February 2023). Distribution of clinic numbers held in intervention CHs is shown in S1 Figure: twelve CHs had no clinics, and, among the other 23 CHs, 16 were pharmacist-led. Where delivered, most CHs had one or two clinics. Only two intervention CHs were paid an incentive fee for achieving staff vaccination rates above 70%.

For the NHS perspective, there was no significant difference between control and intervention arms in unadjusted and adjusted NHS use and costs (see S3 Tables and S4 Table for further detail).

### Outcomes

Differences in percentages of staff vaccinated are reported in Table 3. Compared to CHs in the control arm, the adjusted percentage of staff vaccinated in CHs in the intervention arm was non-significantly (p=0.572) higher on average by 1.8% (95% CI: -6.0%, 10.8%).

There were no significant differences between other outcomes, as reported in Table 4 (all p>0.25). Average sick day totals were similar between arms both for the annual standardised rate and the season adjusted comparison. Agency usage was low irrespective of arm – 60 CHs reported no shifts worked by agency staff. Compared to control CHs, resident death counts were on average lower in the intervention arm in the standardised annual rate and season unadjusted analysis, but higher in the season adjusted analysis.

**Table 4:**
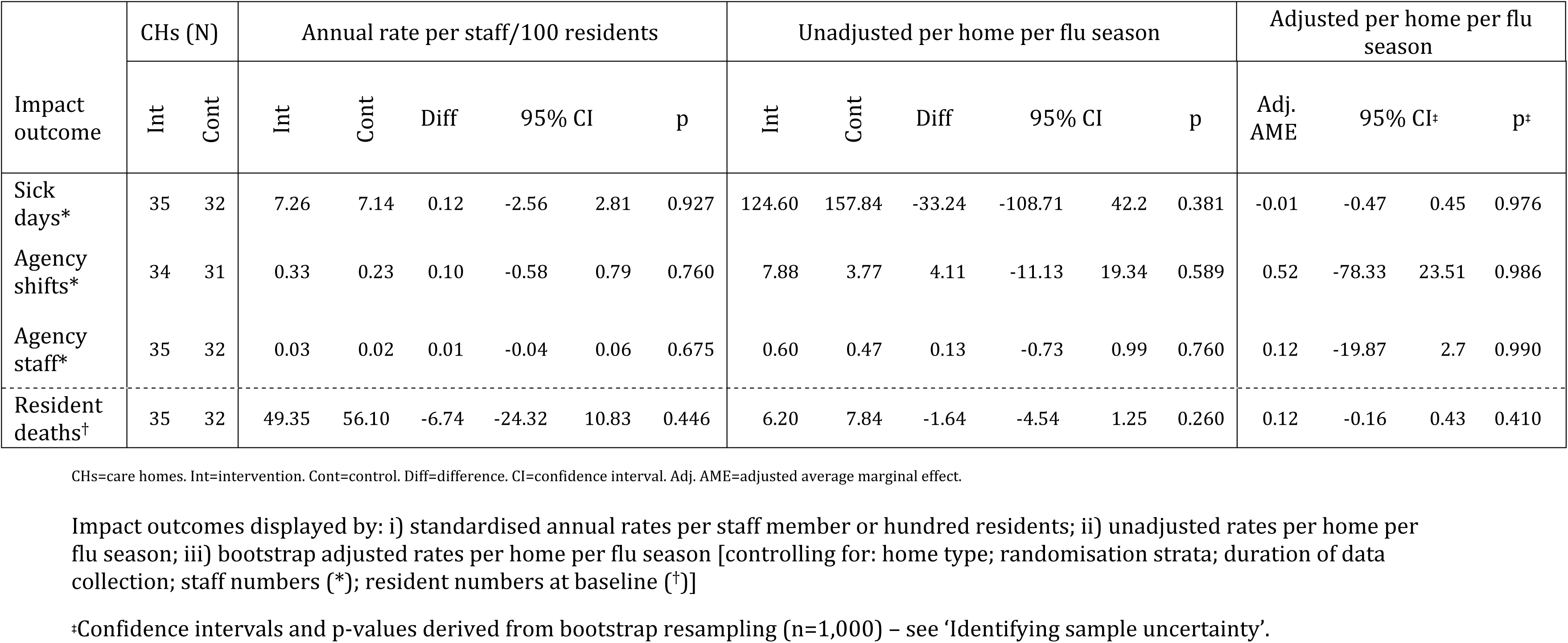
Impact outcomes for base-case for CHs with complete data on intervention costs and resident NHS use.

### Cost-effectiveness of staff vaccination

Cost-effectiveness evidence for FluCare in the base-case analysis is given in Table 3, and Figure 2 and Figure S2. In this setting, there are no established thresholds for determining cost-effectiveness.

**Fig 2:**
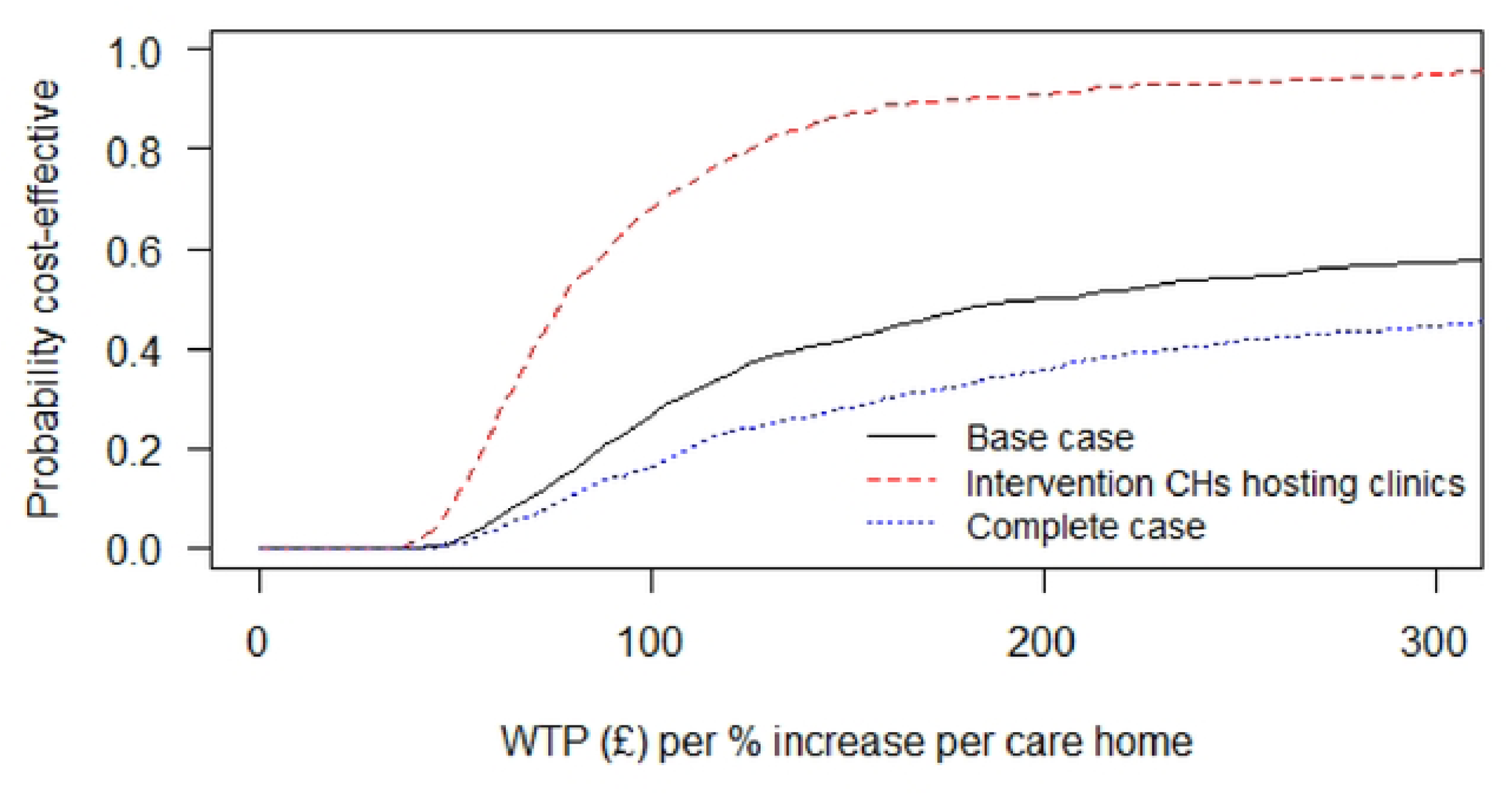
Vaccination funder perspective Cost-effectiveness acceptability curves (CEACs) for: i) base case, ii) intervention CHs hosting clinics and iii) CHs with complete data. CEACs resulting from the: i) base-case analysis; ii) sensitivity analysis (SA) only including intervention care homes (CHs) hosting FluCare vaccination clinics; and iii) SA for CHs with complete data. CEACs show the probability of being cost-effective at different willingness to pay (WTP) thresholds for each vaccination percentage point increase per CH.

CEACs resulting from the: i) base-case analysis; ii) sensitivity analysis (SA) only including intervention care homes (CHs) hosting FluCare vaccination clinics; and iii) SA for CHs with complete data. CEACs show the probability of being cost-effective at different willingness to pay (WTP) thresholds for each vaccination percentage point increase per CH.

From the perspective of the vaccination programme funder, FluCare raises the seasonal vaccination rate at an additional cost of £249 per additional percentage point (PAPP) per CH (adjusted analysis) (see Table 3). From this perspective, the probability of cost-effectiveness exceeds 0.5 for willingness to pay (WTP) values above £204. Results from the NHS perspective are reported in S5 Table.

### Sensitivity analyses

#### Including intervention CHs hosting FluCare vaccination clinics

Only including intervention CHs hosting FluCare vaccination clinics reduced the sample size to 23 intervention homes, whilst the control arm continued with 32 homes. CH characteristics across arms are summarised in S6 Table. Arm differences are similar to the base-case. No agency staff were reported as utilised among the 23 intervention CHs.

Costs between arms are reported in Table 5. From the vaccination funder perspective, mean costs in the intervention arm rose (£1,045 versus £756) compared to the base-case, mainly driven by the greater mean costs of clinic fees. From the NHS perspective, there was no significant difference in costs between arms – further detail is reported in S7 Table, S8 Table and S9 Table.

**Table 5:** Vaccination, intervention and outcome valuation costs/rates per CH per flu season, for SA (intervention homes hosting FluCare clinics; with complete intervention costs and NHS use)

| Cost (£, 21/22)/<br>outcome | CHs (N) |  | Unadjusted per home per flu season |  |  |  |  |  | Adjusted per home per flu season |  |  |  |
| --- | --- | --- | --- | --- | --- | --- | --- | --- | --- | --- | --- | --- |
|  | Int | Cont | Int | Cont | Diff. | 95% CI |  | p | Adj. diff. | 95% CI<sup>‡</sup> |  | p<sup>‡</sup> |
| Vaccine* | 23 | 32 | 212 | 215 | -3 | -99 | 93 | 0.950 | 72 | 3 | 154 | 0.044 |
| Vacc. admin fee* | 23 | 32 | 172 | 174 | -2 | -79 | 76 | 0.969 | 59 | 4 | 125 | 0.040 |
| Promotion | 23 | 32 | 24 | 0 |  |  |  |  |  |  |  |  |
| Clinic fees | 23 | 32 | 563 | 0 |  |  |  |  |  |  |  |  |
| Incentives | 23 | 32 | 74 | 0 |  |  |  |  |  |  |  |  |
| Total* | 23 | 32 | 1,045 | 389 | 656 | 394 | 919 | <0.001 | 805 | 603 | 1,079 | <0.001 |
| % vaccinated * | 23 | 32 | 36.4 | 27.0 | 9.5 | -2.1 | 21.0 | 0.107 | 10.1 | 0.9 | 21.9 | 0.018 |
| Funder ICER: £/% |  |  |  |  | 69 |  |  |  | 79 |  |  |  |
| Funder ICER: £/person |  |  |  |  | 111 |  |  |  | 126 |  |  |  |

CHs=care homes. Int=intervention. Cont=control. Diff=difference. CI=confidence interval. Adj. diff.=adjusted difference.
ICER=incremental cost-effectiveness ratio.

Compared to the control arm, the adjusted percentage of staff vaccinated in the intervention arm was significantly (p=0.018) higher on average by 10.1% (95% CI: 0.9%, 21.9%; adjusted analysis, Table 5): beyond a five-fold increase on the base-case. Other outcomes are reported in Table 6: compared to the base-case, there was minimal evidence of a difference in staff sick days and resident mortality, but there did appear to be a greater reduction in the use of agency staff in the intervention arm (as above, no agency staff were reported as utilised in these intervention CHs).

**Table 6:**
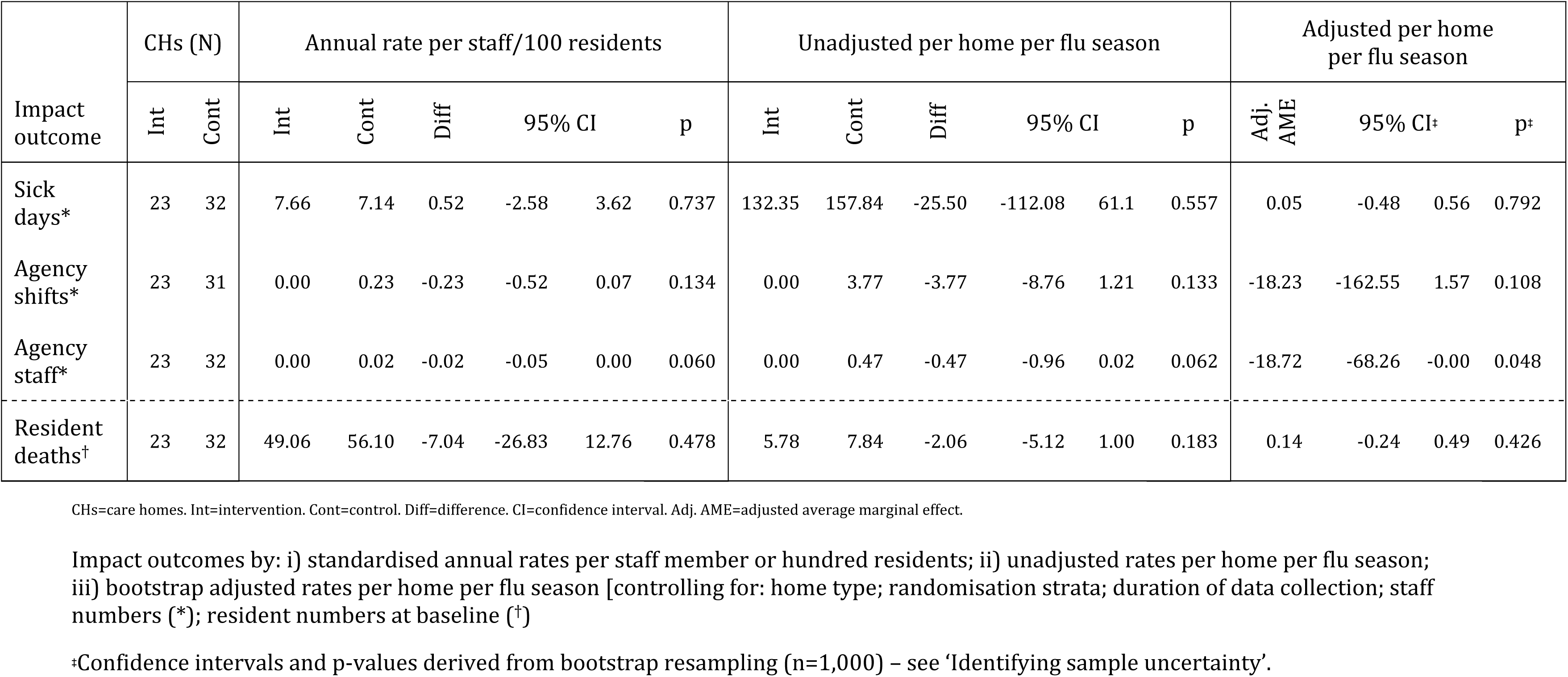
Impact outcomes for sensitivity analysis restricted to intervention homes hosting FluCare clinics, along with complete data on intervention costs and NHS use.

From the funder perspective, the cost PAPP per CH reduces to £79 (adjusted analysis; Table 5), around a third of the base-case. Probability of cost-effectiveness increases more quickly than the base-case, and plateaus at a higher value (above 0.91 from a WTP of £200). See S9 Table for the cost PAPP from the NHS perspective.

#### Complete case analysis

Detail of the complete case analysis is provided in S10 Table and S11 Table. Excluding CHs with partially missing data reduces the sample size to 29 CHs in each arm. From the funder perspective, compared to the base-case, with similar costs but a lower proportion of staff vaccinated (adjusted comparisons: 0.5% versus 1.8%) causes the cost PAPP per CH to rise to £780 (S11 Table). The probability of cost-effectiveness is similar until around a WTP value of £50, and then deviates to being around 0.14 lower (see Fig 2).

For the NHS perspective, see S11 Table for incremental total costs and S2 Fig for associated cost effectiveness acceptability curves.

#### Only including i) direct and bank staff and ii) only direct staff

The impact of restricting staff considered to a) those directly employed or bank staff (so, excluding agency and volunteers), and b) directly employed staff (excluding all other categories), for the i) base-case and ii) sensitivity analysis where intervention CHs are only included if they hosted FluCare vaccination clinics is explored in S12 Table, S13 Table, S14 Table and S15 Table. Associated impacts are mostly clearly seen on the CEACs shown in S3 Fig and S4 Fig: compared to including all staff, cost-effectiveness improves the most when only directly employed staff are considered (excluding bank, agency and volunteers), and to a lesser extent when only agency and volunteers are excluded (numbers within these categories are low – see e.g., Table 2); differences are larger in the base-case than the sensitivity analysis.

### Secondary analyses

#### Valuing impacts on sick days and utilisation of agency staff

Estimated values to CHs of sick days and utilisation of agency staff are reported within Table 4 and S5 Table. Differences between arms on these outcomes were minimal, and so, consequently, differences between them in terms of their value is also minimal.

#### Valuing mortality differences as quality adjusted life years (QALYs)

Accrued QALY differences between arms were small given small mortality differences between them. See Tables A and B in S3 Text for detail.

#### Impact of using staff level derived vaccination effectiveness

This analysis only considers the funder’s perspective: compared to the main analyses, the ICER for the base-case is higher (£360 versus £249 PAPP per CH) and very similar to the sensitivity analysis where only intervention CHs hosting FluCare vaccination clinics are included (£72 versus £79 PAPP per CH). See Table A in S4 Text for further detail.

#### Costing FluCare clinics based on standard payment rates

Detail of alternative costings of FluCare clinics is reported in S5 Text. Only in the case of GPs were FluCare clinic delivery costs greater than the FluCare clinic fee if reimbursed according to typical hourly rates.

## Discussion

This is the first EE of an RCT aiming to improve the health of CH residents in England through staff flu vaccination, benefitting from a CCA to compare impacts across a relatively wide range of domains. The base case analysis indicated that FluCare led a to only a small increase (1.8%) in average CH vaccination rate. However, a pre-specified sensitivity analysis restricted to intervention CHs that hosted FluCare vaccination clinics found a significant (p=0.018) average increase of 10.1%, exceeding the 8% intervention increases seen in two previous flu vaccination studies in CHs [19][20].

Hayward et al. [8] found a just over 40% intervention increase among staff – however, their study had less external validity, taking place within a single (large) company, in a pre-Covid context. Our study’s embedded process evaluation found that staff experience during the pandemic negatively impacted attitudes towards flu vaccination (affecting FluCare too) [45] – for example, mandatory CH staff Covid vaccination undermining autonomy increased resistance to flu vaccination. The broader process evaluation [26] also suggests our results were generally impacted by late delivery of the intervention - two to three months into the flu season. We expect this to have had multiple impacts: it will have been more difficult to deliver clinics due to limited availability of flu vaccines, with less time to do so, and staff less invested in being vaccinated as they perceived the flu season to be almost over, or, may already have been vaccinated. At an organisational level, a CH manager’s motivation to support clinic implementation and opportunity to work towards financial incentives may have also been reduced. Additionally, there was less time in which to accrue evidence of impact on other outcomes – consistent with finding little difference between other outcomes such as resident mortality and staff sick days. However, earlier initiation of FluCare will need to be balanced against evidence of flu vaccination effectiveness waning during the season [46], contributing to the general flu vaccination programme rollout being delayed until October [47].

Cost-effectiveness from the perspective of the programme funder depends on the WTP for improvements in proportions of CH staff vaccinated – we are unaware of commonly agreed values. However, when only including CHs hosting FluCare vaccination clinics in the intervention arm, the probability of being cost-effective quickly increases over 0.5 from a WTP of £80 PAPP per CH, and plateaus at around 0.91 for a WTP of £200 PAPP per CH. When only including CHs hosting FluCare vaccination clinics in the intervention arm, vaccination rate is increased *and* small cost-savings are made: i.e. FluCare was ‘dominant’ and so is (by definition) cost-effective. However, there was a lot of uncertainty – at a WTP of 0, the probability of being cost-effective is only 0.55. Process evaluation evidence suggests it would be detrimental to restrict the FluCare intervention to only consisting of FluCare clinics – however, earlier delivery and more clinics might well be expected to positively impact on cost-effectiveness and allow more time to positively impact other outcomes [26]. Further, from the sensitivity analyses that excluded various types of staff (from among categories of directly employed, bank, agency and voluntary), cost-effectiveness would likely be improved by focusing on directly employed staff. This is consistent from a practical/behavioural stance – for example, the intervention would be expected to less effectively impact agency staff, who may only have a single shift at a given CH.

Differences between arms were minimal when valuing impacts on sick days, utilisation of agency staff and valuing mortality differences as QALYs. We used an approach whereby we costed staff members as a like-for-like replacement, however, staff may not be replaced at all, and/or may be replaced with an agency staff member.

We only found in the case of GP-led vaccination would FluCare clinic delivery costs be greater than the clinic fee if reimbursed according to typical hourly rates. Delivery by community pharmacies will be further facilitated by ‘patient group directions’ allowing registered pharmacy technicians to deliver vaccinations (and other medication) from June 2024 [48]. This may further reduce the cost of pharmacy-led clinics if reimbursed according to typical hourly rates.

This work has limitations. Late delivery of FluCare clinics limits time in which to accrue evidence of impact on outcomes, such as NHS use by CH residents. Other limitations include the scope of data collected, and much of it being aggregate and consequently less sensitive. For example, we have not more directly measured impact on staff and residents; for example, we could have collected utility data from either group to directly estimate QALYs. However, use of aggregate data was a pragmatic choice to maximise data completeness, informed by the feasibility trial [15]. CH recruitment has been shown to be time-consuming and resource-heavy with recruitment rates as low as 5%, and data collection burden cited as an important consideration for managers when choosing whether to participate [49]. The feasibility study also highlighted the infeasibility of separately recording resource use related to ‘influenza-like illness’ – as in Hayward et al. [8] – within resources available to this trial; this may have been more sensitive to change borne by flu vaccination.

Use of agency staff may have been underreported, with 60 CHs reporting zero shifts worked by agency staff. Discussion with stakeholders suggests use of agency staff is perceived negatively, and so some CHs may have underreported. Of course, low reporting may just reflect challenges of tracking this information for a generally peripatetic workforce, or poor recording and availability of these data. Given potential for biased reporting and challenges of data collection, validity maybe increased by using (mandated) routinely collected data, such as the CQC capacity tracker (which, among other data, collects information on staff numbers and their vaccination status [50]).

Lastly, particularly in terms of resource use and resident outcomes, it is worth noting that flu vaccine effectiveness varies season to season. Therefore, comparison between arms will also have been confounded by the effectiveness of the flu vaccine for this particular season. Consequently, a strength of Hayward et al. [8] is that they explore their intervention effectiveness across two seasons, finding differential impacts in terms of resident outcomes. Thus, we benefit from considering both perspectives to the funder and the NHS – the former is much less likely to be impacted by the particular effectiveness of a given season’s vaccine.

## Conclusions

This is the first EE of an intervention to increase flu vaccination among CH staff in England. Although FluCare led to a small increase (1.8%) in CH staff flu vaccination rate, when only including CHs hosting vaccination clinics in the intervention arm, a significant (p=0.018) increase (10.1%) was found. Cost-effectiveness depends on WTP for increased staff vaccination, but cost PAPP per CH improved from £249 to £79 when only considering intervention CHs hosting vaccination clinics. Ability to impact outcomes may have been reduced by initiating FluCare two to three months into flu season – for example, reducing numbers of onsite vaccination clinics. We suggest evaluating FluCare in future flu seasons, initiating it earlier in the season.

## Acknowledgements

We are grateful for the hard work and support of the FluCare team and our PPI group (Saiqa Ahmed, Alison Bryant, Hilary Garrett, Saima Gul, Keith Holt and Hilary Tetlow). We also thank members of the FluCare Project Steering Committee and Data Management and Ethics Committee. Chris Pearson (HC-One Group) has been particularly supportive of FluCare. We acknowledge Norfolk and Waveney Integrated Care System and the UEA who have hosted/sponsored respectively. Both institutions provided unwavering support from inception onwards. The views expressed are those of the authors and not necessarily those of the NIHR or the DHSC.

## Data availability statement

Requests for FluCare trial dataset access should be made to Dr Amrish Patel and/or. Post-trial requests will be reviewed by Chief Investigators (and representatives from the Trial Management Group where available) and Sponsor. Requests will be reviewed to ensure no conflict with FluCare trial objectives (pre-publication of FluCare findings) or funder constraints. All data requests will be subject to a data-sharing agreement with the University of East Anglia (UEA).

Code is available upon reasonable request by emailing APW

## Supporting information captions

### Text file captions

S1 Text: Deviations to the health economic analysis plan (HEAP).

S2 Text: Further detail, assumptions, source and unit costs for FluCare

S3 Text: Secondary analysis: Valuing mortality differences as QALYs

S4 Text: Secondary analysis: Using staff level effectiveness analysis

S5 Text: Secondary analysis: Alternative FluCare clinic costings.

### Figure captions

S1 Fig: Distribution of intervention care home (CH) numbers split by the number of onsite clinics delivered by pharmacist or general practitioner (GP).

S2 Fig: NHS perspective CEACs for: i) base case, ii) intervention CHs with FluCare clinics and iii) CHs with complete data.

S3 Fig: Vaccination funder perspective CEACs for: i)base-case and SA (intervention CHs with clinics) by staff category.

S4 Fig: NHS perspective CEACs for the base-case and SA (intervention CHs with clinics) by staff category.

S5 Fig: Cost-effectiveness planes for base-case analysis (left column) and sensitivity analysis where intervention CHs are restricted to those holding FluCare clinics (right column), for funder (top row) and NHS perspective (bottom row).

### Table captions

S3 Table: Resident NHS use for base-case with complete intervention costs and complete NHS use.

S4 Table: Costs of resident NHS use for base-case with complete intervention costs and complete NHS use.

S5 Table: Vaccination, intervention and outcome valuation costs/rates for base-case with complete intervention costs and complete NHS use.

S6 Table: Description of care homes included within base-case analysis where intervention homes had vaccination clinics (complete intervention costs and complete NHS).

S7 Table: Resident NHS use for base-case where intervention homes had vaccination clinics with complete intervention costs and complete NHS use.

S8 Table: Costs of resident NHS use for base-case where intervention homes had vaccination clinics with complete intervention costs and complete NHS use.

S9 Table: Vaccination, intervention and outcome valuation costs/rates for base-case where intervention homes had vaccination clinics with complete intervention costs and complete NHS use.

S10 Table: Description of care homes included within complete case analysis (complete intervention costs and complete NHS).

S11 Table: Vaccination, intervention and outcome valuation costs, for complete case data (no missing data imputation) with complete intervention costs and complete NHS use.

S12 Table: Vaccination, intervention and outcome valuation costs, for base-case with complete intervention costs and complete NHS, *only* including directly employed and bank staff (e.g. excluding agency and volunteers).

S13 Table: Vaccination, intervention and outcome valuation costs, for base-case with complete intervention costs and complete NHS, ONLY including directly employed staff (e.g. excluding bank, agency and volunteers).

S14 Table: Vaccination, intervention and outcome valuation costs, for base-case where intervention homes had vaccination clinics alongside complete intervention costs and complete NHS, ONLY including directly employed and bank staff (e.g. excluding agency and volunteers).

S15 Table: Vaccination, intervention and outcome valuation costs, for base-case where intervention homes had vaccination clinics alongside complete intervention costs and complete NHS, ONLY including directly employed staff (e.g. excluding bank, agency and volunteers).

S16 Table: Unit costs and assumptions for secondary analyses costing FluCare clinics based on timings recorded in the vaccine provider log. £, 2021/22.

## References

1. UK Health Security Agency. Surveillance of influenza and other seasonal respiratory viruses in the UK, winter 2022 to 2023. 2023; Available from: https://www.gov.uk/government/statistics/annual-flu-reports/surveillance-of-influenza-and-other-seasonal-respiratory-viruses-in-the-uk-winter-2022-to-2023

2. Matias G, Taylor RJ, Haguinet F, Schuck-Paim C, Lustig RL, Fleming DM. Modelling estimates of age-specific influenza-related hospitalisation and mortality in the United Kingdom. BMC Public Health. 2016;16:481.

3. Ellis SE, Coffey CS, Mitchel EF, Dittus RS, Griffin MR. Influenza- and respiratory syncytial virus-associated morbidity and mortality in the nursing home population. J Am Geriatr Soc. 2003;51:761–7.

4. Gallagher N, Johnston J, Crookshanks H, Nugent C, Irvine N. Characteristics of respiratory outbreaks in care homes during four influenza seasons, 2011-2015. J Hosp Infect. 2018;99:175–80.

5. Goodwin K, Viboud C, Simonsen L. Antibody response to influenza vaccination in the elderly: A quantitative review. Vaccine. 2006;24:1159–69.

6. Franklin B, Hochlaf D. An Economic Analysis of Flu Vaccination [Internet]. London: International Longevity Centre—UK; 2018. Available from: https://ilcuk.org.uk/wp-content/uploads/2018/07/An_economic_analysis_of_flu_vaccination_-_ILC-UK.pdf

7. Carman WF, Elder AG, Wallace LA, McAulay K, Walker A, Murray GD, et al. Effects of influenza vaccination of health-care workers on mortality of elderly people in long-term care: A randomised controlled trial. Lancet. 2000;355:93–7.

8. Hayward AC, Harling R, Wetten S, Johnson AM, Munro S, Smedley J, et al. Effectiveness of an influenza vaccine programme for care home staff to prevent death, morbidity, and health service use among residents: Cluster randomised controlled trial. BMJ. 2006;333:1241.

9. Lemaitre M, Meret T, Rothan-Tondeur M, Belmin J, Lejonc JL, Luquel L, et al. Effect of influenza vaccination of nursing home staff on mortality of residents: A cluster-randomized trial. J Am Geriatr Soc. 2009;57:1580–6.

10. Public Health England. Guidelines for PHE health protection teams on the management of outbreaks of influenza-like illness (ILI) in care homes [Internet]. 2020. Available from: https://assets.publishing.service.gov.uk/media/5fa3f65b8fa8f5788e288cd4/Guidelines_for_the_management_of_outbreaks_of_influenza-like_illness_in_care_homes_05_11_2020.pdf

11. Public Health England. Flu and flu vaccination 2020/21: A toolkit for care homes [Internet]. 2019. Available from: https://www.england.nhs.uk/south/wp-content/uploads/sites/6/2020/09/Care-Home-toolkit.docx-Final-2021.pdf

12. National Institute for Health and Care Excellence. Flu vaccination: Increasing uptake (NICE guideline) [Internet]. Public Health England,; 2018. Available from: https://www.nice.org.uk/guidance/ng103/resources/flu-vaccination-increasing-uptake-pdf-66141536272837

13. NHS England and NHS Improvement. Social care: Guidance for workforce flu vaccination. 2019.

14. Department of Health & Social Care. Adult social care in England, monthly statistics: quarterly update to February 2026 [Internet]. 2026. Available from: Adult social care provider statistics, England: quarterly update to February 2026 - GOV.UK

15. Wright D, Patel A, Blacklock J, Bion V, Birt L, Bryant T et al. FluCare: Results from a randomised feasibility study of a complex intervention to increase care home staff influenza vaccination rates. Archives of Clinical and Biomedical Research. 2024 Jul 2;8:273–290. doi: 10.26502/acbr.50170410

16. Wright D, Blacklock J, Bion V, Birt L, Clark A, Griffiths AW et al. Effectiveness of a theory-informed intervention to increase care home staff influenza vaccination rates: A cluster randomised controlled trial. Journal of Public Health. 2025 Jun;47(2):246–257. Epub 2025 Mar 30. doi: 10.1093/pubmed/fdaf023

17. National Institute for Health and Care Excellence. Flu vaccination: Increasing uptake - evidence reviews for increasing uptake in health and social care staff. NICE guideline NG103 [Internet]. 2018. Available from: https://www.nice.org.uk/guidance/ng103/evidence/4-increasing-flu-vaccination-uptake-in-health-and-social-care-staff-pdf-6532083617

18. Burls A, Jordan R, Barton P, Olowokure B, Wake B, Albon E, et al. Vaccinating healthcare workers against influenza to protect the vulnerable–is it a good use of healthcare resources? A systematic review of the evidence and an economic evaluation. Vaccine [Internet]. 2006;24:4212–21. Available from: https://www.sciencedirect.com/science/article/pii/S0264410X05012946?via%3Dihu b

19. Chen H, Ng S, King ME, Fong C, Ng W, Szeto K, et al. Promotion of seasonal influenza vaccination among staff in residential care homes for elderly in hong kong. Healthcare infection [Internet]. 2010;15:121—125. Available from: https://europepmc.org/articles/PMC7129256

20. Lorini C, Ierardi F, Gatteschi C, Galletti G, Collini F, Peracca L., et al. Promoting influenza vaccination among staff of nursing homes according to behavioral insights: Analyzing the choice architecture during a nudge-based intervention. Vaccines (Basel) [Internet]. 2020;8:600. Available from: https://europepmc.org/articles/PMC7129256

21. Dey P, Halder S, Collins S, Benons L, Woodman C. Promoting uptake of influenza vaccination among health care workers: A randomized controlled trial. J Public Health Med. 2001;4:346–8.

22. Carman WF, Elder AG, Wallace LA, McAulay K, Walker A, Murray GD, et al. Effects of influenza vaccination of health-care workers on mortality of elderly people in long-term care: A randomised controlled trial. The Lancet [Internet]. 2000;355:93–7. Available from: https://www.sciencedirect.com/science/article/pii/S0140673699051909

23. Pereira M, Williams S, Restrick L, Cullinan P, Hopkinson NS. Healthcare worker influenza vaccination and sickness absence–an ecological study. Clinical Medicine. 2017;17:484.

24. Ofori SK, Hung YW, Schwind JS, Diallo K, Babatunde D, Nwaobi SO, et al. Economic evaluations of interventions against influenza at workplaces: Systematic review. Occup Med (Lond). 2022;72:70–80.

25. Patel A, Sims E, Blacklock J, Birt L, Bion V, Clark A, et al. Cluster randomised control trial protocol for estimating the effectiveness and cost-effectiveness of a complex intervention to increase care home staff influenza vaccination rates compared to usual practice (FLUCARE). Trials. 2022;23:989.

26. Katangwe-Chigamba T, Alsaif F, Anyiam-Osigwe A, Bion V, Clark A, Garrett H, et al. Process evaluation of the flucare cluster randomised controlled trial: assessing the implementation of a behaviour change intervention to increase influenza vaccination uptake among care home staff in England. BMC Health Serv Res. 2025;25(1):1118. doi: 10.1186/s12913-025-13298-0.

27. Tudor Edwards R, McIntosh E. Applied Health Economics for Public Health Practice and Research [Internet]. Oxford University Press; 2019. Available from: 10.1093/med/9780198737483.001.0001

28. Mounier-Jack S, Bell S, Chantler T, Edwards A, Yarwood J, Gilbert D, et al. Organisational factors affecting performance in delivering influenza vaccination to staff in NHS acute hospital trusts in england: A qualitative study. Vaccine [Internet]. 2020;38:3079–85. Available from: https://www.ncbi.nlm.nih.gov/pmc/articles/PMC7090903/

29. Sand KL, Lynn J, Bardenheier B, Seow H, Nace DA. Increasing influenza immunization for long-term care facility staff using quality improvement. J Am Geriatr Soc. 2007;55:1741–7.

30. Allan S, Forder J. Care markets in england: Lessons from research [Internet]. University of Kent, Kent: Personal Social Services Research Unit; 2012. Available from: https://www.pssru.ac.uk/pub/dp2815.pdf

31. Johnston SS, Rousculp MD, Palmer LA, Chu B-C, Mahadevia PJ, Nichol KL. Employees’ willingness to pay to prevent influenza. The American Journal of Managed Care. 2010;16:e205–14.

32. NHS England. Community pharmacy advanced service specification: Seasonal influenza vaccination 1 September 2024 - 31 March 2025: Version 2.0 [Internet]. 2024. Available from: https://www.england.nhs.uk/wp-content/uploads/2017/08/R008-11-PRN00996iii-cp-seasonal-influenza-vaccination-as-spec-aw-2425-june-2024.pdf

33. NHS England. Seasonal influenza vaccination programme 2022/23 [Internet]. 2022. Available from: https://www.england.nhs.uk/london/wp-content/uploads/sites/8/2022/08/B1772_Seasonal-influenza-vaccination-prog-22-23-ESS-for-GPs_August-2022-.pdf

34. Patel A, Scott S, Griffiths AW, & Wright, D. Development of a behaviour change intervention to increase care home staff influenza vaccination uptake. International Journal of Nursing Studies Advances. 2025;9:100387. 10.1016/J.IJNSA.2025.100387

35. Hoffmann T, Glasziou P, Boutron I, Milne R, Perera R, Moher D, et al. Better reporting of interventions: Template for intervention description and replication (TIDieR) checklist and guide. BMJ. 2014;348.

36. NHS Business Services Authority. Prescription cost analysis – England – 2021/22 [Internet]. 2022. Available from: https://www.nhsbsa.nhs.uk/statistical-collections/prescription-cost-analysis-england/prescription-cost-analysis-england-202122

37. NHS Business Services Authority. NHS electronic drug tariff: Part VIC - Advanced Services (Pharmacy and Appliance Contractors)(England) [Internet]. 2023. Available from: https://www.drugtariff.nhsbsa.nhs.uk/#/00841024-DC/DD00840781/Part%20VIC%20-%20Advanced%20Services%20(Pharmacy%20and%20Appliance%20Contractors)(England)

38. Jones K, Weatherly H, Birch S., Castelli A, Chalkley M, Dargan A, et al. Unit Costs of Health and Social Care 2021 manual. Curtis L, Burns A, editors. Canterbury: Personal Social Services Research Unit, University of Kent & Centre for Health Economics (University of York); 2023.

39. Drummond MF, Sculpher MJ, Claxton K, Stoddart GL, Torrance GW. Methods for the Economic Evaluation of Health Care Programmes. 4th ed. Oxford University Press, Oxford; 2015.

40. Leeper TJ. Margins: Marginal effects for model objects. 2024.

41. Efron B, Tibshirani RJ. An introduction to the bootstrap. 1st ed. London: Chapman; Hall/CRC; 1994.

42. Black WC. The CE plane: A graphic representation of cost-effectiveness. Medical decision making: an international journal of the Society for Medical Decision Making. 1990;10:212–4.

43. Fenwick E, O’Brien BJ, Briggs A. Cost-effectiveness acceptability curves–facts, fallacies and frequently asked questions. Health economics. 2004;13:405–15.

44. Logan PA, Horne JC, Allen F, Armstrong SJ, Clark AB, Conroy S, et al. A multidomain decision support tool to prevent falls in older people: The FinCH cluster RCT. Health Technol Assess [Internet]. 2022;26:1–136. Available from: https://www.ncbi.nlm.nih.gov/books/n/ukhta2609/pdf/

45. Anyiam-Osigwe A, Katangwe-Chigamba T, Scott S, Seeley C, Patel A, Sims EJ et al. A psychosocial critique of the consequences of the COVID-19 pandemic on UK care home staff attitudes to the flu vaccination: A qualitative longitudinal study. Vaccines. 2024 Dec 20;12(12):1437. doi: 10.3390/vaccines12121437

46. Joint Committee on Vaccination and Immunisation. Minutes of the joint committee on vaccination and immunisation [Internet]. 2023. Available from: https://app.box.com/s/iddfb4ppwkmtjusir2tc/file/1440465623207

47. Department of Health & Social Care. National flu immunisation programme 2024 to 2025 letter [Internet]. 2024. Available from: https://www.gov.uk/government/publications/national-flu-immunisation-programme-plan-2024-to-2025/national-flu-immunisation-programme-2024-to-2025-letter

48. Department of Health & Social Care. Consultation outcome: Proposal to enable pharmacy technicians to supply and administer medicines using patient group directions: Consultation response [Internet]. 2024. Available from: https://www.gov.uk/government/consultations/proposal-for-the-use-of-patient-group-directions-by-pharmacy-technicians/outcome/proposal-to-enable-pharmacy-technicians-to-supply-and-administer-medicines-using-patient-group-directions-consultation-response#next-steps

49. Ellwood A, Airlie J, Cicero R, Cundill B, Ellard DR, Farrin A, et al. Recruiting care homes to a randomised controlled trial. Trials, 2018;19(1): 535. 10.1186/s13063-018-2915-x

50. Department of Health and Social Care. Formal notice of a mandate for all adult social care (ASC) providers: Information required by the secretary of state for health and social care from CQC-regulated ASC providers, under section 277A of the health and social care act 2012, as inserted by section 99 of the health and care act 2022 [Internet]. 2024. Available from: https://www.gov.uk/government/publications/adult-social-care-provider-information-provisions-formal-notice/formal-notice-of-a-mandate-for-all-adult-social-care-providers

